# Discussion of Mental Illness and Mental Health By NBA Players on Twitter

**DOI:** 10.1101/2020.10.19.20215681

**Authors:** Shaan Kamal, Osama El-Gabalawy, Shahan Kamal, Mohammed H. Malik

## Abstract

In 2019, the National Basketball Association (NBA) expanded its mental health rules to include mandating that each team have at least one mental health professional on their full-time staff and to retain a licensed psychiatrist to assist when needed. In this work, we investigate the NBA players’ discussion of mental health using historical data from players’ public Twitter accounts. All current and former NBA players with Twitter accounts were identified, and each of their last 800 tweets were scraped, yielding 920,000 tweets. A list of search terms derived from the DSM5 diagnoses was then created and used to search all of the nearly one million tweets. In this work, we present the most common search terms used to identify tweets about mental health, present the change in month-by-month tweets about mental health, and identify the impact of players discussing their own mental health struggles on their box score statistics before and after their first tweet discussing their own mental health struggles.

## Introduction

Although sports psychiatry is not a new area within psychiatry, athlete mental health has become a more prevalent topic of discussion as many successful, world renowned athletes have discussed their own mental health struggles.^1–3^ Despite the ongoing discussions of mental health in this population, there is little in the way of literature on the topic or any research quantifying such disclosures by athletes. Furthermore, the literature on psychiatric treatment of athletes in professional organizations is also limited.^4^ In 2020, the National Basketball Association (NBA) expanded its mental health policy to include mandating that each team have at least one mental health professional on their full-time staff, to retain a licensed psychiatrist to assist when needed, maintain confidentiality, and provide services that are culturally competent.^5^ These measures were a response to increasing player discussions of mental health on social media platforms.^5^ In January of 2021, the NBA enacted policy requiring teams to provide telehealth services in light of the ongoing COVID-19 pandemic for mental health. In this work, we investigate the NBA players’ discussion of mental health using historical data from players’ public Twitter accounts.

## Methods

All current and former NBA players with Twitter accounts were identified using BasketballReference, and each of their last 800 tweets were scraped in June 2020 via the official Twitter API. This included players’ own tweets as well as retweets from other accounts. A list of search terms derived from mental health terms and DSM5 diagnoses was used to search all of the collected tweets (Supplemental Figure 1). All tweets were read to determine if players were disclosing any personal mental health struggles. We present the most common search terms used to identify tweets about mental health, the monthly change in such tweets, and identify the impact of players’ disclosure on their box score statistics.

## Results

In total, 1374 active Twitter accounts were found for current and former NBA players, with a total of 902,454 tweets collected. Using the designated search terms, 1,293 tweets were found to contain at least one mental health term. Of the players with a Twitter account, 508 players (37%) tweeted about mental health. The average number of likes for all collected tweets was 6,484 and for mental health related tweets was 16,533. There is a clear trend of an increasing number of positive tweets each year with a large increase in 2018, coinciding with the first NBA player mental health Twitter disclosure (Figure 1).

**Figure 1:**
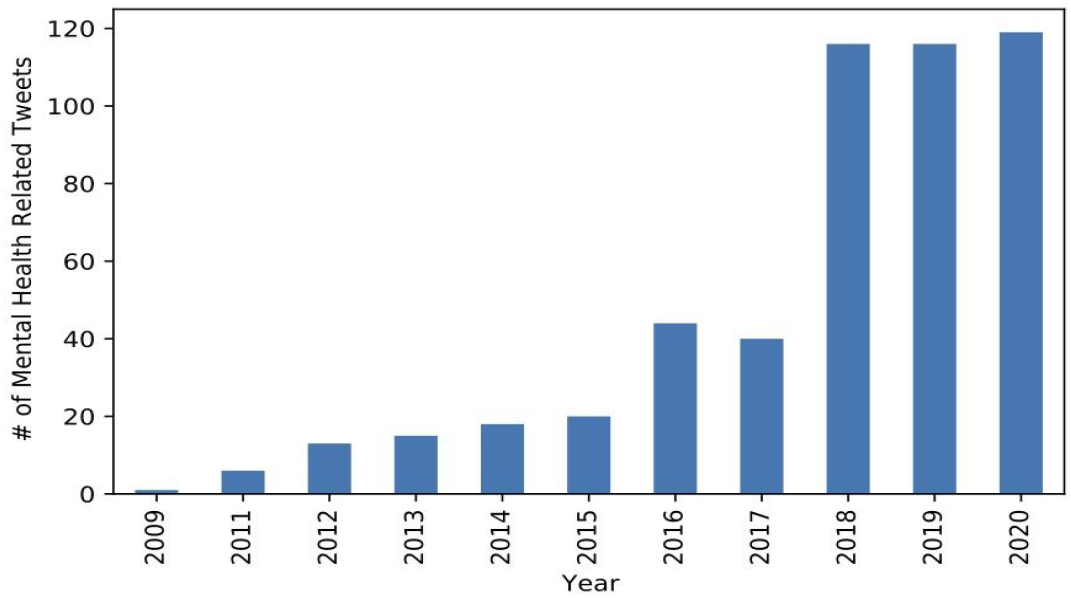
Number of Mental Health Related Tweets by Year

The tweets were then analyzed to determine which search terms were most common within the subset of tweets about mental health (Figure 2). The five most common terms found (mental health, autism, anxiety, suicide, and depression) accounted for 80% of all mental health tweets.

**Figure 2:**
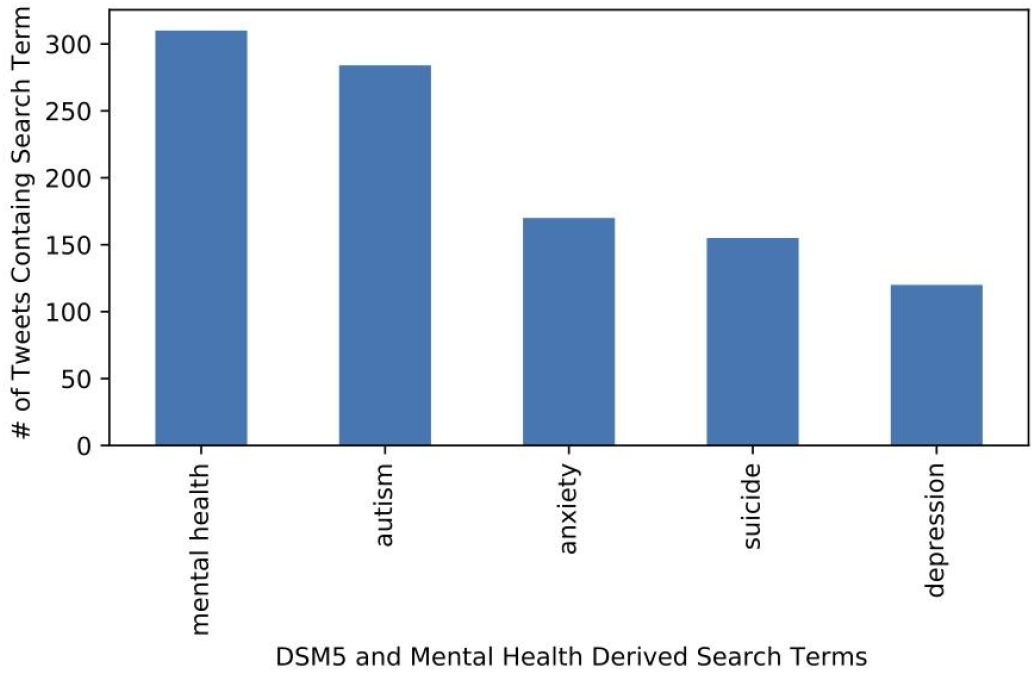
Top Five Most Commonly Found Search Terms

Tweets disclosing personal mental health struggles were used to examine any potential correlation between disclosure and performance. Two players were found to fit this criteria. These players’ performance in the 10 games before and after their disclosure was compared using BasketballReference’s boxscores (Supplemental Figure 2). Both players had better performance in the 10 games after in almost all statistical categories, although both also had increases in personal fouls and turnovers.

## Discussion

Sports psychiatry is a growing field, with increasing literature on psychiatrists working with professional organizations. Twitter has become a leading platform for public figures, such as professional athletes, to voice opinions. This work adds to the literature in showing quantitatively how the discussion amongst professional athletes regarding mental health has changed over time and that tweets about mental health show more engagement with the general public. In addition, there appears to be a positive correlation between disclosure and player performance.

## Data Availability

All data in the manuscript is publicly available from the sources described, our work just provides an analysis for it.

https://www.twitter.com

## Author Contributions

Shaan Kamal and Osama El-Gabalawy both developed the methodology, analyzed results, and wrote and edited the final manuscript. Shahan Kamal and Mohammed Malik assisted in methodology and manuscript review.

## Supplemental Figures

**Supplemental Figure 1:**
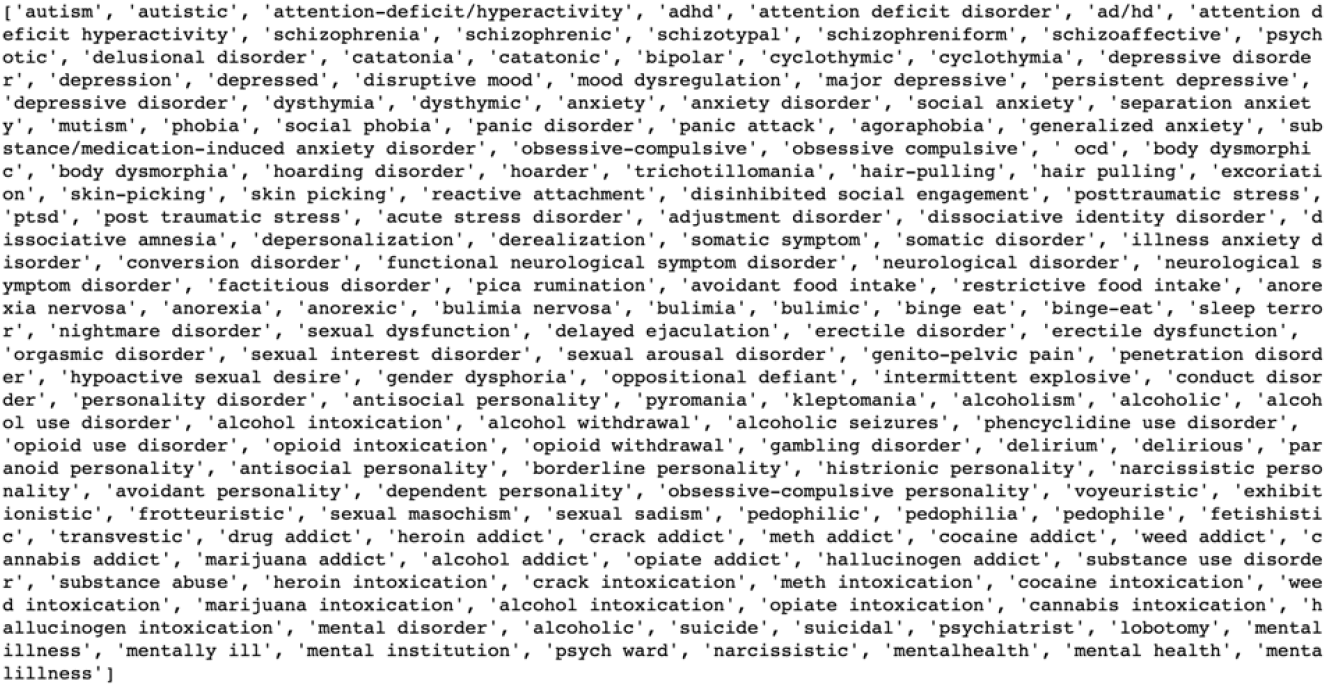
DSM5 and Mental Health Derived Search Terms

**Supplemental Figure 2:**
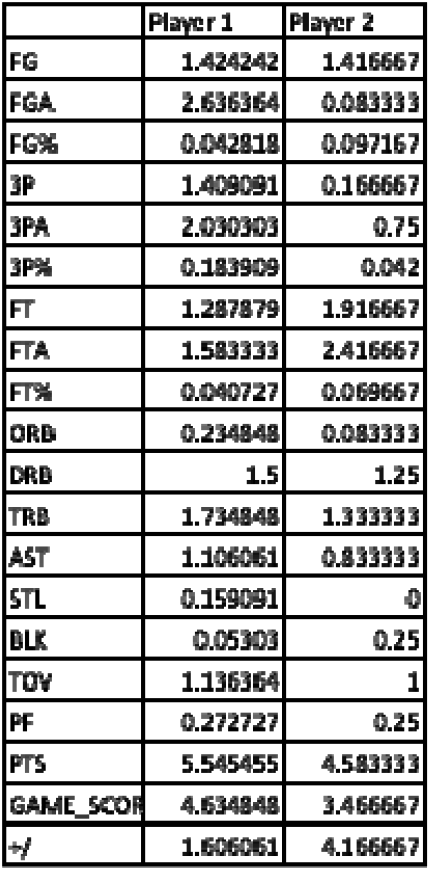
Players’ performance difference in the 10 games after disclosing personal mental health struggles compared to the 10 games afterwards

