## Supplementary figures and images for "Discussion of Mental Illness and Mental Health By NBA Players on Twitter"

### Supplemental Figure 1

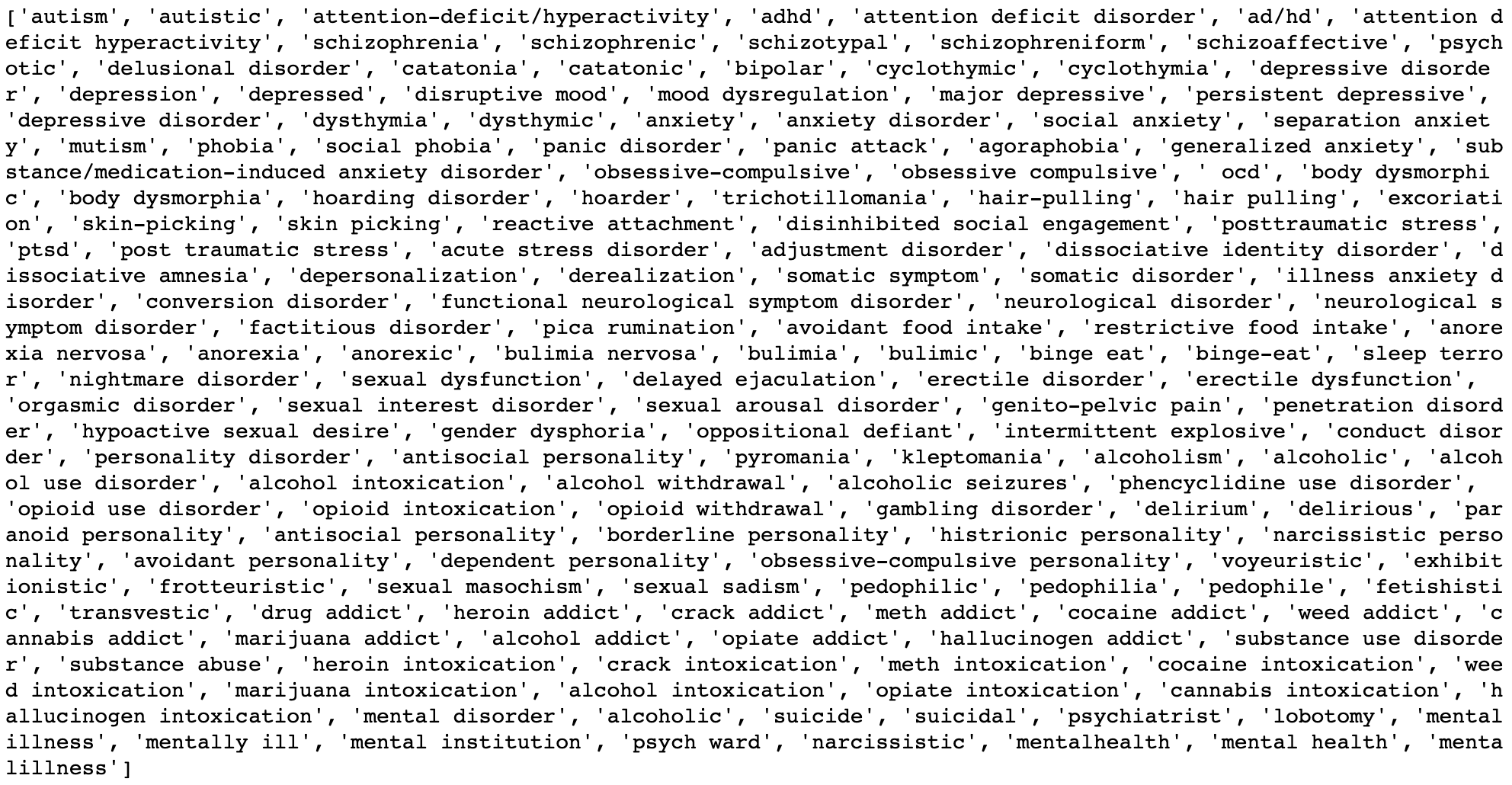

### Supplemental Figure 2

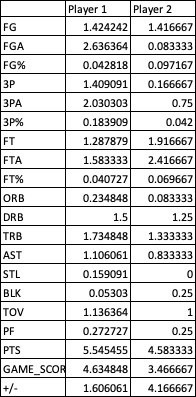
